# A Wearable Fiber-Free Optical Sensor for Continuous Monitoring of Neonatal Cerebral Blood Flow and Oxygenation

**DOI:** 10.1101/2023.09.21.23295914

**Authors:** Xuhui Liu, Mehrana Mohtasebi, Pegah Safavi, Faraneh Fathi, Samaneh Rabienia Haratbar, Li Chen, Jin Chen, Henrietta S. Bada, Lei Chen, Elie G. Abu Jawdeh, Guoqiang Yu

## Abstract

**Impact:**

- The innovative DSCFO technology may serve as a low-cost wearable sensor for continuous bedside monitoring of multiple cerebral hemodynamic parameters in neonatal intensive care units.
- Concurrent DSCFO and DCS measurements of CBF variations in neonatal piglet models generated consistent results.
- No consistent correlation patterns were observed among peripheral and cerebral monitoring parameters in preterm neonates, suggesting the importance of multi-parameter measurements for understanding deep insights of peripheral and cerebral regulations during IH events.
- Integrating and correlating multiple cerebral functional parameters with clinical outcomes may identify biomarkers for prediction and management of IH associated brain injury.

**Background:** Unstable cerebral hemodynamics places preterm infants at high risk of brain injury. We adapted an innovative, fiber-free, wearable diffuse speckle contrast flow-oximetry (DSCFO) device for continuous monitoring of both cerebral blood flow (CBF) and oxygenation in neonatal piglets and preterm infants.

**Methods:** DSCFO uses two small laser diodes as focused-point and a tiny CMOS camera as a high-density two-dimensional detector to detect spontaneous spatial fluctuation of diffuse laser speckles for CBF measurement, and light intensity attenuations for cerebral oxygenation measurement. The DSCFO was first validated against the established diffuse correlation spectroscopy (DCS) in neonatal piglets and then utilized for continuous CBF and oxygenation monitoring in preterm infants during intermittent hypoxemia (IH) events.

**Results:** Consistent results between the DSCFO and DCS measurements of CBF variations in neonatal piglets were observed. IH events induced fluctuations in CBF, cerebral oxygenation, and peripheral cardiorespiratory vitals in preterm infants. However, no consistent correlation patterns were observed among peripheral and cerebral monitoring parameters.

**Conclusions:** This pilot study demonstrated the feasibility of DSCFO technology to serve as a low-cost wearable sensor for continuous monitoring of multiple cerebral hemodynamic parameters. The results suggested the importance of multi-parameter measurements for understanding deep insights of peripheral and cerebral regulations.

## 1. Introduction

An estimated 15 million preterm infants are born annually in the world ^1^. Preterm infants are at the highest risk for developing brain injury and subsequent long-term neurodevelopmental impairment. Multiple factors contribute to brain injury and poor outcomes in preterm infants including their hemodynamic instability, impaired cerebral autoregulation, and frequent intermittent hypoxemia (IH) events ^2–5^. IH is defined as episodic drops in arterial blood oxygen saturation (SpO_2_), measured by peripheral pulse oximeters. Mounting evidence derived from both animal models and clinical studies suggests that IH is associated with brain injury and poor outcomes such as increased inflammation, impaired growth, retinopathy of prematurity, and neurodevelopmental impairment ^6,7^. A considerable challenge in neonatal intensive care thus far has been the reliable direct monitoring of brain hemodynamics. Many groups currently utilize peripheral cardiorespiratory and hemodynamic monitoring as surrogate indicators of cerebral blood flow and oxygenation in preterm infants. However, peripheral monitoring may not consistently offer a reliable indicator of brain health. A few groups have tested near-infrared spectroscopy (NIRS) in preterm infants with IH and found fluctuations in cerebral oxygenation. However, it is difficult to establish predictive thresholds for treatment decisions via NIRS alone due to its large intra and inter-patient variability of cerebral oxygenation ^8–10^. Consequently, there is a growing interest in alternative methods that can measure both cerebral blood flow (CBF) and cerebral oxygenation since these two together are likely more sensitive biomarkers of brain health than one single parameter alone. Moreover, integrating/correlating temporal/spatial variation patterns of physiological and cerebral parameters during IH with clinical outcomes may generate robust biomarkers for assessing and predicting IH associated consequences ^11,12^.

Conventional cerebral hemodynamics monitoring modalities such as magnetic resonance imaging (MRI) and transcranial Doppler (TCD) ultrasound are not suitable for continuous monitoring of CBF in preterm infants due to either motion artifact (MRI) or the excessive size of the probe (TCD) ^13^. A variety of wearable techniques have been developed recently for continuous bedside cerebral monitoring. Among them, NIRS has been widely used as noninvasive bedside techniques for continuous monitoring of cerebral blood oxygenation ^14,15^. Conventional NIRS measures light intensity attenuations by tissue absorption and scattering at multiple wavelengths to calculate changes in oxy- and deoxy-hemoglobin concentrations ([HbO_2_] and [Hb]) ^16,17^. The recently developed wearable/wireless NIRS systems facilitate the continuous monitoring of subjects in a comfortable manner although they measure cerebral blood oxygenation only ^18–20^. Near-infrared (NIR) diffuse correlation spectroscopy (DCS) is a relatively new technology that uses coherent NIR light and single-photon-counting avalanche photodiodes (APDs) to detect temporal fluctuations of diffuse laser speckles resulting from red blood cell motions in the brain (i.e., CBF) ^21–24^. DCS has been used for continuous monitoring of CBF variations in preterm infants ^22,25^. Moreover, DCS has been combined with NIRS into hybrid instruments for simultaneous measurements of CBF and cerebral oxygenation ^17,26,27^. While effective, DCS utilizes large, expensive, long-coherence lasers and discrete single-photon-counting APDs for CBF measurements, which limits the spatial-temporal resolution and increases instrumentation dimension and cost. Moreover, the hybrid fiber-optic probe coupling of NIRS/DCS sources and detectors is difficult to install on the small heads of neonates for continuous and longitudinal cerebral monitoring. In addition, rigid and fragile optical fibers used for source-detector (S-D) couplings are sensitive to motion artifact and significantly constrain subject’s movement ^25,28–30^.

To overcome these challenges, we have recently developed a NIR diffuse speckle contrast flow oximetry (DSCFO) that provides a low-cost, fiber-free, wearable, and compact approach for continuous monitoring blood flow and oxygenation variations in freely behaving subjects ^31–34^. DSCFO uses two small laser diodes as focused-point sources for deep tissue penetration and a tiny CMOS camera as a high-density two-dimensional (2D) detector array to detect spontaneous spatial fluctuations of diffuse laser speckles for blood flow detection, and light intensity attenuations at different wavelengths for blood oxygenation measurement. Thousands of parallel pixels on the CMOS sensor significantly improve the temporal-spatial resolution and reduce the cost and dimension of the probe/device. The connections between the wearable probe and DSCFO device are all flexible electrical wires (i.e., fiber free), enabling continuous monitoring of tissue blood flow and oxygenation variations in freely behaving subjects. The sensitivity and accuracy of DSCFO for blood flow and oxygenation measurements have been previously verified against established NIRS and DCS technologies in tissue-simulating phantoms, human forearm muscles during arterial cuff-occlusion, and mouse brains under anesthesia and during freely behavior tests with hypercapnia challenges ^31–35^.

In the present study, the DSCFO system was scaled up and adapted for continuous monitoring of CBF, [HbO_2_], and [Hb] in larger heads of neonatal piglets and human preterm infants, which are more difficult to penetrate through than small rodents. Continuous measurements of cerebral hemodynamic variations were conducted concurrently by the DSCFO and dual-wavelength DCS devices in neonatal piglet models of hypercapnia (resulting in CBF increase) and transient global ischemia and asphyxia (resulting in CBF decrease). Neonatal piglets are chosen because their brain and development as well as head size are analogous to human neonates, which provides an opportunity to assess whether DSCFO detects cerebral hemodynamic alterations under ischemic/hypoxic stress in newborn subjects. The DSCFO was then used in the neonatal intensive care unit (NICU) to continuously monitor CBF and cerebral oxygenation variations during IH in preterm infants. Results were compared to other physiological parameters for understanding pathophysiological impacts of IH on premature brains.

## 2. Methods

Our innovative DSCFO and established DCS technologies were used in this study for comparisons. The details of DSCFO and DCS techniques can be found in the **Supplementary Materials**. Four neonatal piglets were studied using the DSCFO and DCS devices during hypercapnia, transient global ischemia, and asphyxia. Animal experimental protocols have been approved by the Institutional Animal Care and Use Committee (IACUC) at the University of Kentucky. The DSCFO was then tested in five preterm infants to continuously monitor variations in CBF and cerebral oxygenation during IH events in the NICU. Other physiological parameters such as heart rate (HR), respiration rate (RR), SpO_2_, and/or end-tidal CO_2_ (EtCO_2_), were also continuously recorded by commercial devices in the clinic. Human experimental protocols have been approved by the Institutional Review Board (IRB) at the University of Kentucky.

### 2.1 Animal Experimental Protocols

Four female neonatal Yorkshire piglets (postnatal age: 4 days) were purchased from the Swine Center at the University of Kentucky and housed in a designated room for animal experiments. **Fig. 1** shows study purposes, instrument/probe configurations, and study protocols in all animal experiments. **Fig. 2** illustrates experimental setup for continuous monitoring of cerebral hemodynamics in neonatal piglets using different DSCFO and DCS probes. Before experiments, animals were kept in an animal incubator with proper temperature and humidity and fed with milk replacer (Brithright, Ralco) every 4 hours. The piglet was fasted for at least 4 hours before anesthesia to avoid reflux of stomach contents. The animal was induced with 5% Isoflurane inhalation in a customized anesthesia chamber, and then intubated with a pediatric endotracheal tube (internal diameter: 3 mm) and maintained with 1.5% - 2% Isoflurane. The anesthetized animal was placed prone with its head secured on a customized stereotaxic frame (**Fig. 2a**). Animal’s body was covered with an electrical heated blanket under the monitoring of a rectal thermometer to avoid hypothermia.

**Fig. 1:**
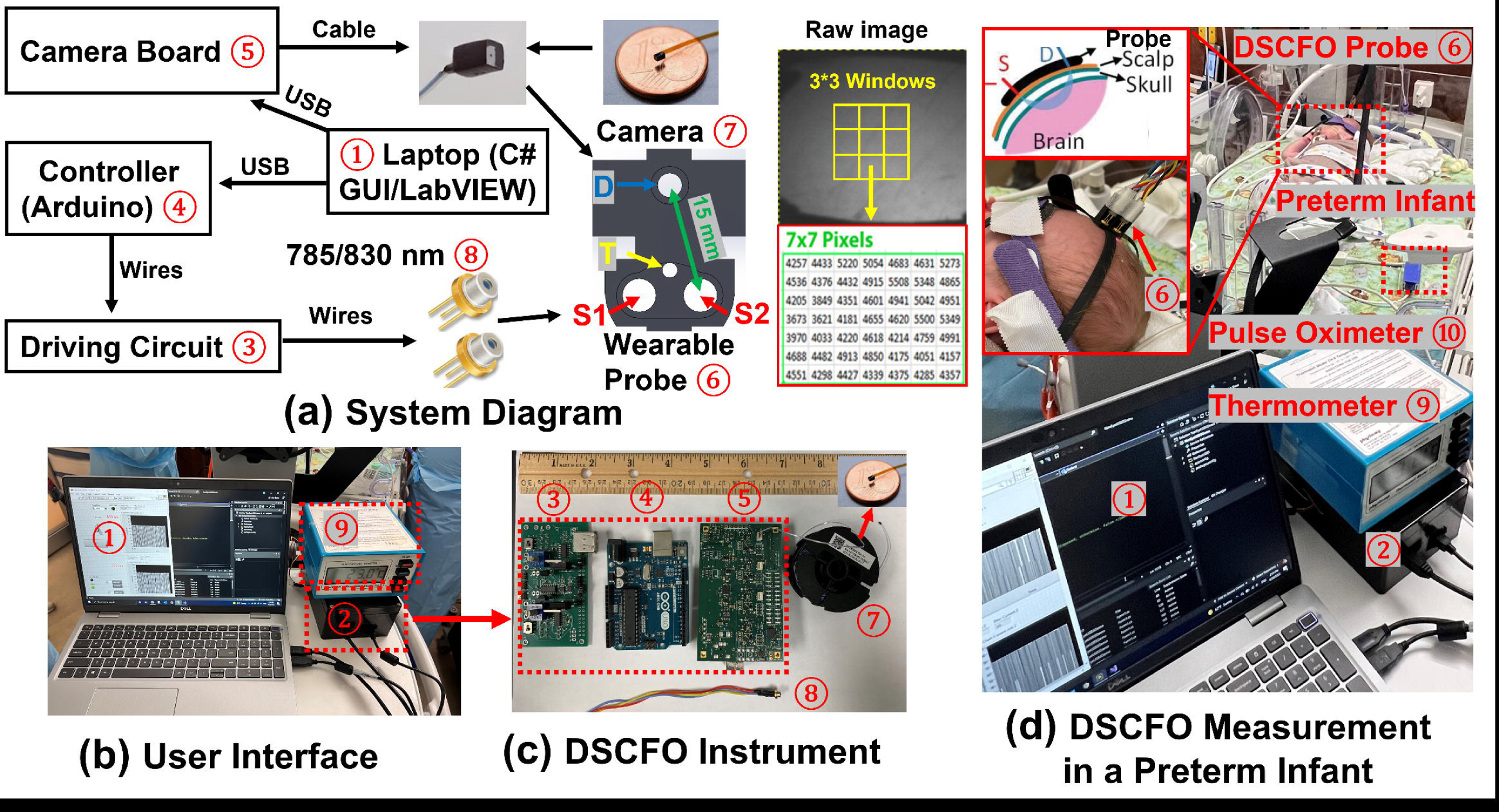
Animal experimental protocols and corresponding optical measurements with specific probes.

**Fig. 2:**
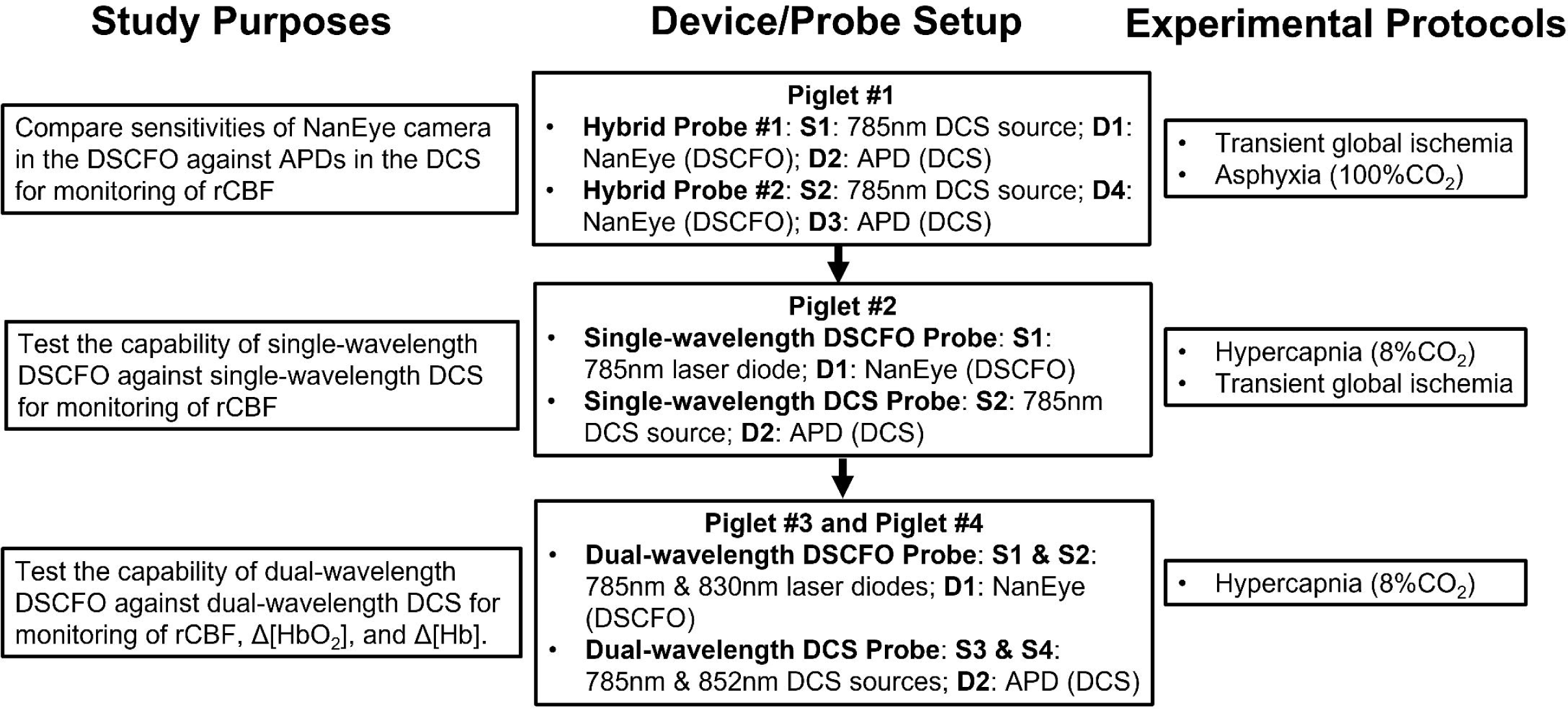
Continuous monitoring of cerebral hemodynamics in neonatal piglets using different DSCFO and DCS probes. **(a)** Experimental setup for continuous monitoring of multiple physiological parameters in a neonatal piglet. **(b)** Two hybrid probes (Probe #1 and Probe #2) installed on the intact skull of right and left hemispheres of Piglet #1, respectively. Each probe consisted of one DCS source (S1 or S2: 785 nm long-coherence laser, CrystaLaser) and two detectors (D1 or D4: NanEye cameras; D2 or D3: APDs in DCS). **(c)** A single-wavelength DSCFO and a single-wavelength DCS probes installed on the scalp of right and left hemispheres of Piglet #2, respectively. The DSCFO probe consisted of one source (S1: 785 nm laser diode, Thorlabs) and one detector (D1: NanEye camera). The DCS probe had one source (S2: 785 nm long-coherence laser, CrystaLaser) and one detector (D2: APD). **(d)** A dual-wavelength DSCFO and a dual-wavelength DCS probes installed on the scalp of right and left hemispheres of Piglet #3 and Piglet #4, respectively. The DCSFO probe consisted of two sources (S1: 785 nm laser diode; S2: 830 nm laser diode; Thorlabs) and one detector (D1: NanEye camera). The DCS probe had two sources (S3: 785 nm long-coherence laser; S4: 852 nm long-coherence laser, CrystaLaser) and one detector (D2: APD).

*Continuous monitoring of rCBF during transient global ischemia and asphyxia (100%CO_2_) in Piglet #1.* Piglet #1 was used to test the sensitivity of NanEye cameras in the DSCFO against the APDs in the DCS for continuous monitoring of rCBF during transient global ischemia and asphyxia (**Fig. 2b**). Under anesthesia, the animal’s scalp was retracted to expose the skull. Two hybrid probes (Probe #1 and Probe #2) were glued on the skull of right and left hemispheres, respectively. Each of these two probes consisted of one DCS source (S1 or S2: 785 nm long-coherence laser, CrystaLaser) and two detectors (D1 or D4: NanEye cameras; D2 or D3: APDs in DCS). The sampling rates of two hybrid probes were 2 Hz.

The transient global ischemia and asphyxia were induced following our established methods in neonatal piglets ^36^. Briefly, after hairs on the ventral cervical area were shaved with a clipper and cleaned with hair cream, the animal was laid supine, and a 5-cm midline incision was made in the cervical area. The skin was opened with four customized surgical hooks for better exposure of the surgical area. The underlying tissue was split with blunt dissection until both left and right common carotid arteries (CCAs) were exposed. A braided 6-0 sterile surgical suture was wrapped around each CCA, and a loose knot was made on each side without disrupting blood flow to the brain. The animal was then laid prone, and the scalp was surgically opened. Probe #1 and Probe #2 were glued on the exposed skull. After a baseline measurement for ∼7.5 minutes, the knot on the left CCA was tightened to occlude the left CCA for ∼7 minutes, followed by tightening the right knot for ∼2 minutes to induce transient global ischemia. The right and left knots were then released sequentially, allowing the restoration of blood flow to the brain. After recovering from transient global ischemia, the piglet received asphyxia through inhalation of 100%CO_2_ via a Y-shape connector with regulator valves before sacrifice.

*Continuous monitoring of rCBF during hypercapnia (8%CO_2_) and transient global ischemia in Piglet #2.* Piglet #2 was used to test the capability of single-wavelength DSCFO against single-wavelength DCS for continuous monitoring of rCBF during hypercapnia and transient global ischemia (**Fig. 2c**). A single-wavelength DSCFO probe and a single-wavelength DCS probe were glued on the intact scalps of right and left hemispheres, respectively. The fiber-free DCSFO probe consisted of one source (S1: 785 nm laser diode, Thorlabs) and one detector (D1: NanEye camera). The fiber-optic DCS probe had one source (S2: 785 nm long-coherence laser, CrystaLaser) and one detector (D2: APD). The sampling rates of two probes were 2 Hz.

The hypercapnia and transient global ischemia were induced following our established methods ^37^. After ∼10 minutes baseline recording of rCBF by the DSCFO/DCS probes, the mixed gas of 8%CO_2_/92%O_2_ was administered through a Y-shape connector for ∼10 minutes. The CO_2_ was then stopped, and DSCFO/DCS measurements lasted for another ∼11 minutes to record the recovery of rCBF. The transient global ischemia was then applied as described above. Briefly, after a 3-minute baseline DSCFO/DCS measurement, the left CCA was occluded for ∼7 minutes, followed by occluding the right CCA for ∼2 minutes to induce the transient global ischemia. The right and left CCAs were then released sequentially to allow cerebral reperfusion.

*Simultaneous measurements of rCBF,* Δ*[HbO_2_], and* Δ*[Hb] during hypercapnia (8%CO_2_) in Piglet #3 and Piglet #4.* Piglet #3 and Piglet #4 were used to test the capability of dual-wavelength DSCFO against dual-wavelength DCS for continuous monitoring of rCBF, Δ[HbO_2_], and Δ[Hb] during hypercapnia (**Fig. 2d**). A dual-wavelength DSCFO probe and a dual-wavelength DCS probe were glued on the intact scalps of right and left hemispheres, respectively. The fiber-free DCSFO probe consisted of two sources (S1: 785 nm laser diode; S2: 830 nm laser diode; Thorlabs) and one detector (D1: NanEye camera). The fiber-optic DCS probe had two sources (S3: 785 nm long-coherence laser; S4: 852 nm long-coherence laser, CrystaLaser) and one detector (D2: APD). The sampling rates of two probes were 2 Hz.

The hypercapnia was generated following our established methods ^37^. After ∼5 minutes baseline recording of rCBF by the dual-wavelength DSCFO/DCS probes, the mixed gas of 8%CO_2_/92%O_2_ was administered through a Y-shape connector for ∼10 minutes. The CO_2_ was then stopped, and the measurements continued for another ∼5 minutes to record the recovery of rCBF.

### 2.2 Human Experimental Protocols

Five preterm infants were recruited from the Kentucky Children’s Hospital NICU. Informed parental consents were obtained for all subjects. **Table 1** lists the demographic and baseline characteristics of the 5 enrolled subjects. All infants had no severe intraventricular hemorrhage (IVH) as determined by routine head ultrasounds. All patients had brain MRI prior to NICU discharge with no severe injury found.

**Table 1:**
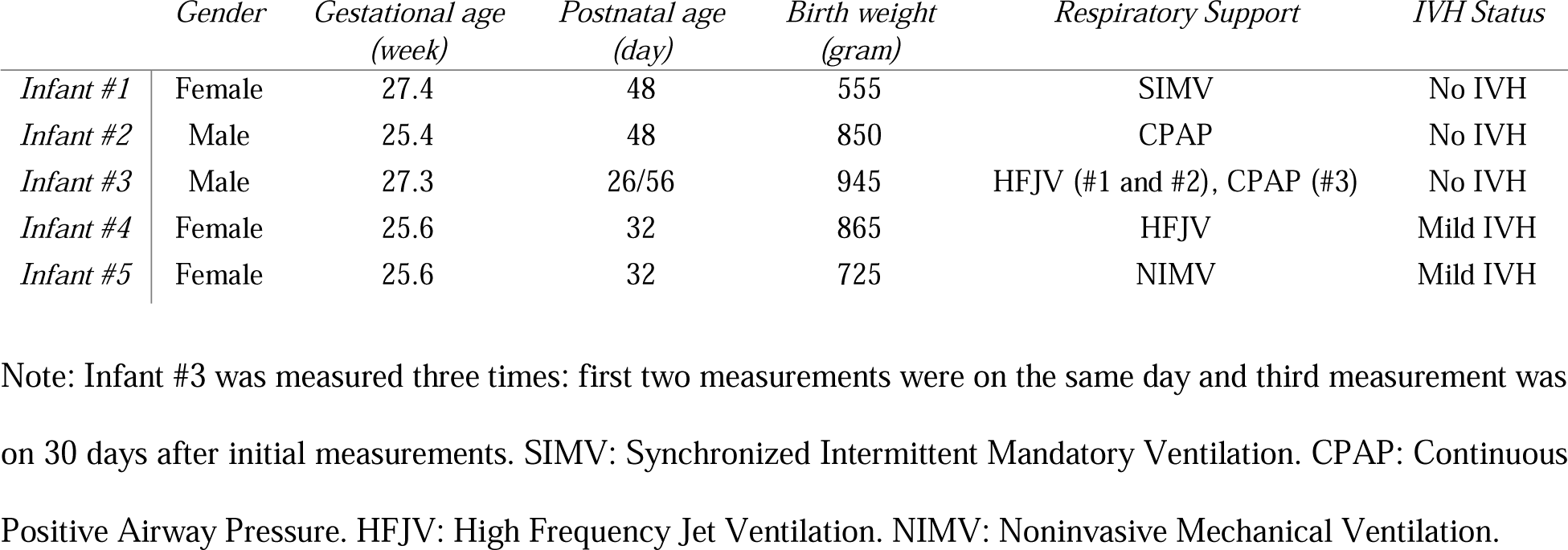
Demographics and baseline characteristics of subjects.

Cerebral hemodynamic changes (rCBF, Δ[HbO_2_], Δ[Hb]) were continuously monitored by the DSCFO probe (**Supplementary Fig. 1**) with a S-D distance of 15 mm and at a sampling rate of 2 Hz over ∼6 minutes. In total, seven measurements were taken from five infants including 3 repeated measurements in Infant #3. No motion artifacts were observed during those measurements. In addition, SpO_2_ was continuously measured by a high-resolution pulse oximeter (sampling rate: 1 Hz, Masimo). The HR, RR, and EtCO_2_ were continuously recorded by a peripheral cardiorespiratory monitoring system (IntelliVue MX800, Philips).

### 2.3 Statistical Analysis

For concurrent DCS and DSCFO measurements in neonatal piglets, linear regression models and Pearson’s correlation coefficients were used to investigate the correlations between rCBF, Δ[HbO_2_], and Δ[Hb] measurements from two different probes. For continuous DSCFO measurements in preterm infants, Pearson’s correlation coefficients were calculated to investigate the correlations between different physiological parameters. A value of p < 0.05 was considered significant. The Pearson’s correlation coefficient r <= 0.3 suggested a weak correlation; 0.3 < r < 0.7 suggested a moderate correlation; and r >= 0.7 suggested a strong correlation. The analyses were performed using MATLAB 2022.

## 3. Results

### 3.1 Animal Experimental Results

*The NanEye camera in DSCFO enabled the detection of rCBF variations during transient global ischemia and asphyxia (**Fig. 3**).* **Fig. 3a** and **Fig. 3b** show time-course rCBF changes during transient global ischemia in Piglet #1, measured concurrently by the two hybrid probes (**Fig. 2b**). Measured rCBF declined from the baseline ⍰ during right CCA occlusion ⍰, and further decreased to the minimal values in both hemispheres during bilateral ligation ⍰ (right hemisphere: 37% and 24%; left hemisphere: 32% and 21%; measured by NanEye and APD, respectively). Note that right CCA occlusion ⍰ might induce significant activation of sympathetic nervous system, leading to rapid blood flow increases through the contralateral carotid artery, and compensating the CBF loss in the ipsilateral hemisphere with collateral flow through the Circle of Willis, as suggested by the rCBF spikes in both hemispheres ^38^. After sequentially releasing the left ligation ⍰ and right ligation ⍰, rCBF then gradually recovered till the end of the measurements. These changes meet physiological expectations. Importantly, significant strong correlations between the concurrent NanEye and APD measurements of rCBF were observed in both right hemisphere (r = 0.91, p < 10^-5^, slope = 1.23; **Fig. 3a**) and left hemisphere (r = 0.90, p < 10^-5^, slope = 1.06; **Fig. 3b**).

**Fig. 3:**
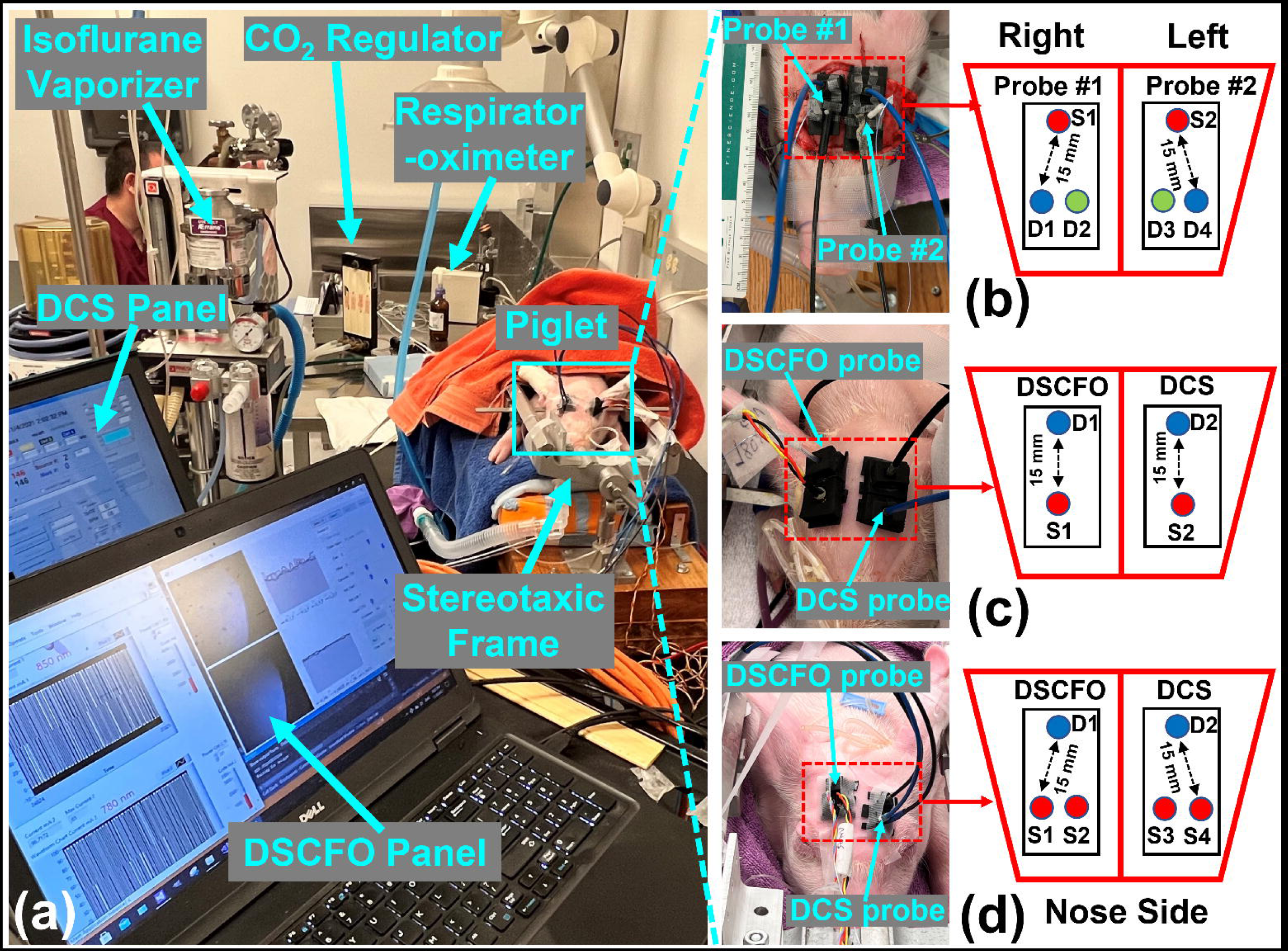
rCBF measurement results from Piglet #1 during transient global ischemia and asphyxia. **(a)** and **(b)** Time-course changes in rCBF during transient global ischemia were measured concurrently by two hybrid probes placed on right (Probe #1) and left (Probe #2) hemispheres. ⍰ Baseline; ⍰ Right CCA ligation; ⍰ Bilateral ligation; ⍰ Releasing left ligation; ⍰ Releasing both ligations & recovery. Linear regression results from Probe #1: y = 1.23x – 15.62, r = 0.91 and from Probe #2: y = 1.06x – 2.04, r = 0.90. Here, x and y represent NanEye and APD measurements, respectively. **(c)** and **(d)** Time-course changes in rCBF during asphyxia (100%CO_2_) measured concurrently by two hybrid probes. Linear regression results from Probe #1: y = 0.87x + 8.94, r = 0.92 and from Probe #2: y = 0.88x + 11.47, r = 0.89.

**Fig. 3c** and **Fig. 3d** show time-course changes in rCBF during global asphyxia (100%CO_2_). At the end of measurements, rCBF dropped to minimal values in both hemispheres (< 8% measured by NanEye; < 5% measured by APD). Significant strong correlations were also observed between the concurrent NanEye and APD measurements of rCBF in both right hemisphere (r = 0.92, p < 10^-5^, slope = 0.87; **Fig. 3c**) and left hemisphere (r = 0.89, p < 10^-5^, slope = 0.88; **Fig. 3d**).

Note that the two hybrid probes consisting of both NanEye and APD were placed on each of the two hemispheres (**Fig. 2b**). Therefore, it is expected to observe strong correlations between concurrent DSCFO and DCS measurements on both hemispheres during sequential CCA ligations (**Fig. 3a** and **Fig. 3b**) and during asphyxia (**Fig. 3c** and **Fig. 3d**).

Single-wavelength DSCFO and single-wavelength DCS measurements of rCBF during hypercapnia and transient global ischemia generated expected results (**Fig. 4**). **Fig. 4a** shows time-course changes in rCBF during 8%CO_2_ inhalation in Piglet #2, measured concurrently by the DSCFO (on right hemisphere) and DCS (on left hemisphere) probes (**Fig. 2c**). As expected, global elevations of rCBF were detected by the DSCFO (+21%) and DCS (+32%) during CO_2_ inhalation. Significant strong correlations between the DSCFO and DCS measurements of rCBF were observed (r = 0.80, p < 10^-5^, slope = 1.31; **Fig. 4a**).

**Fig. 4:**
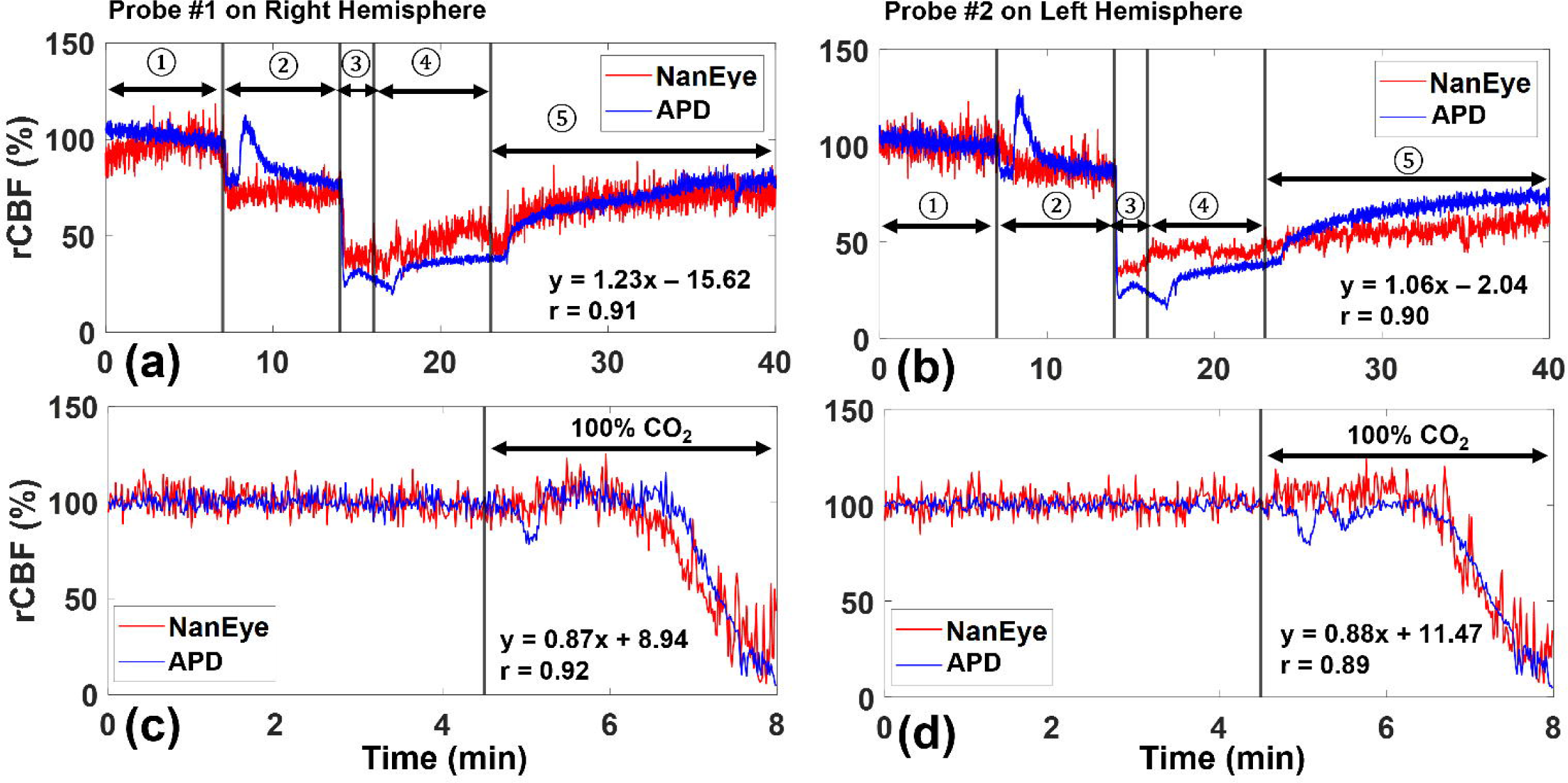
rCBF measurements results from Piglet #2 during hypercapnia and transient global cerebral ischemia. **(a)** Time-course changes in rCBF during 8%CO_2_ inhalation, measured concurrently by the single-wavelength DSCFO probe on right hemisphere and single-wavelength DCS probe on left hemisphere. Linear regression results between the two measurements: y = 1.31x – 27.22, r = 0.80. Here, x and y represent single-wavelength DSCFO and single-wavelength DCS measurements, respectively. **(b)** Time-course changes in rCBF during transient global ischemia measured concurrently by the two probes. ⍰: Baseline; ⍰: Left CCA ligation; ⍰: Bilateral ligation; ⍰: Releasing right ligation; ⍰: Releasing both ligations & recovery.

**Fig. 4b** shows time-course changes in rCBF during transient global ischemia in Piglet #2. During left CCA occlusion ⍰, rCBF on the left hemisphere measured by the DCS rapidly reduced to 60% and then slowly recovered to 75% of the baseline. rCBF on the right hemisphere measured by the DSCFO gradually decreased to 67% of the baseline. These rCBF changes are reasonable because the collateral circulation through the Circle of Willis compensated the left hemisphere during left CCA ligation ⍰, leading to different rCBF responses on left and right hemispheres. During bilateral ligations ⍰, both devices detected further decreases in rCBF, reaching 41% in DSCFO and 35% in DCS of their baseline values, respectively. After sequentially releasing the right ligation ⍰ and left ligation ⍰, the rCBF then gradually recovered towards baseline until the end of the measurement.

Note that hypercapnia induced consistent global changes in rCBF on both hemispheres whereas sequential CCA ligations resulted in different regional changes in rCBF on different hemispheres (**Fig. 4b**). Therefore, it is expected to observe strong correlations between the DSCFO (on right hemisphere) and DCS (on left hemisphere) measurements (**Fig. 2c**) during hypercapnia (**Fig. 4a**) but weaker correlations (r = −0.63) during sequential CCA ligations (**Fig. 4b**).

*Dual-wavelength DSCFO and dual-wavelength DCS measurements of rCBF,* Δ*[HbO_2_] and* Δ[Hb] during hypercapnia generated consistent results (**Fig. 5**). **Fig. 5a** and **Fig. 5b** show the time-course variations in rCBF during 8%CO_2_ inhalation in Piglet #3 and Piglet #4, measured by the DSCFO and DCS probes at 785 nm (**Fig. 2d**). As expected, both probes detected global increases in rCBF during hypercapnia. Significant moderate correlations between the two measurements of rCBF were observed in Piglet #3 (r = 0.62, p < 10^-5^, slope = 1.20; **Fig. 5b**) and in Piglet #4 (r = 0.61, p < 10^-5^, slope = 1.16; **Fig. 5b**). **Fig. 5c** and **Fig. 5d** show the time-course changes in Δ[HbO_2_] during 8%CO_2_ inhalation in Piglet #3 and Piglet #4 respectively, measured by DSCFO and DCS probes at two wavelengths (**Fig. 3d**). Significant strong correlation between the two measurements of Δ[HbO_2_] was observed in Piglet #3 (r = 0.75, p < 10^-5^, slope = 1.24; **Fig. 5c**) and significant moderate correlation in Piglet #4 (r = 0.32, p < 10^-5^, slope = 0.80; **Fig. 5d**). **Fig. 5e** and **Fig. 5f** show the time-course changes in Δ[Hb] during 8%CO_2_ inhalation in Piglet #3 and Piglet #4 respectively, measured by DSCFO and DCS. Weak correlations between the two measurements of Δ[Hb] were observed in Piglet #3 (r = 0.1, p < 10^-5^, slope = 0.21; **Fig. 5e**) and Piglet #4 (r = 0.24, p < 10^-5^, slope = −0.35; **Fig. 5f**).

**Fig. 5:**
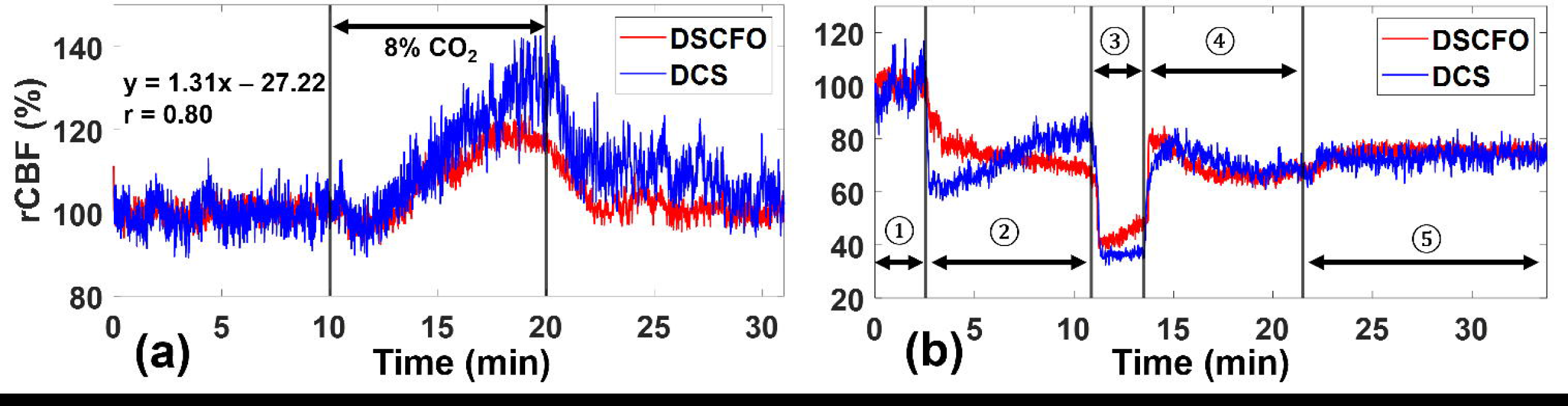
rCBF, Δ[HbO_2_], and Δ[Hb] measurement results from Piglet #3 and Piglet #4 during hypercapnia (8%CO_2_). **(a)** and **(b)** Time-course changes in rCBF during hypercapnia in Piglet #3 and Piglet #4, measured by the dual-wavelength DSCFO and dual-wavelength DCS probes. Linear regression results for Piglet #3 (y = 1.20x + 19.30, r = 0.62) and Piglet #4 (y = 1.16x – 16.45, r = 0.61). Here, x and y represent dual-wavelength DSCFO and dual-wavelength DCS measurements, respectively. **(c)** and **(d)** Time-course changes in Δ[HbO_2_] during hypercapnia in Piglet #3 and Piglet #4, respectively. Linear regression results for Piglet #3 (y = 1.24x + 4.30, r = 0.75) and Piglet #4 (y = 0.80x – 3.10, r = 0.32). **(e)** and **(f)** Time-course changes in Δ[Hb] during hypercapnia in Piglet #3 and Piglet #4, respectively. Linear regression results for Piglet #3 (y = 0.21x – 6.77, r = 0.10) and Piglet #4 (y = −0.35x – 5.22, r = 0.24).

Note that hypercapnia induced consistent global changes in rCBF, Δ[HbO_2_], and Δ[Hb] on both hemispheres. Therefore, it is expected to observe significant correlations between the dual-wavelength DSCFO (on right hemisphere) and dual-wavelength DCS (on left hemisphere) measurements (**Fig. 2d**) during hypercapnia (**Fig. 5**).

### 3.2 Human Experimental Results

Peripheral and regional cerebral regulations during IH events were observed in all 5 preterm infants (see **Fig. 6** and **Supplementary Fig. 2-6**). **Fig. 6a** and **Fig. 6b** show representative results from Infant #1 and Infant #3 (1^st^ measurement), respectively. Data collected from the peripheral cardiorespiratory monitor and pulse oximeter were first interpolated to align with the DSCFO data. Every 10 data points were then averaged to smooth responsive curves. In Infant #1 (**Fig. 6a**), there were two consecutive IH events with SpO_2_ drop to less than 90%. There were increases in HR and RR and a decrease in EtCO_2_ during both events. These peripheral physiological changes resulted in rCBF increases, Δ[HbO_2_] decreases, and Δ[Hb] increases, which started after first IH event and were most noted during the second IH event. In Infant #3 (**Fig. 6b**), HR and RR increases were observed similarly when SpO_2_ dropped down during IH event. Unlike Infant #1, however, rCBF increased ∼20% during the first IH event whereas it decreased during the second event. However, cerebral oxygenation level was maintained at a relatively stable level during the first IH event whereas it presented large variations during the second IH event. The measured skin temperature remained less than 36.8 °C during the measurements for all preterm infants.

**Fig. 6:**
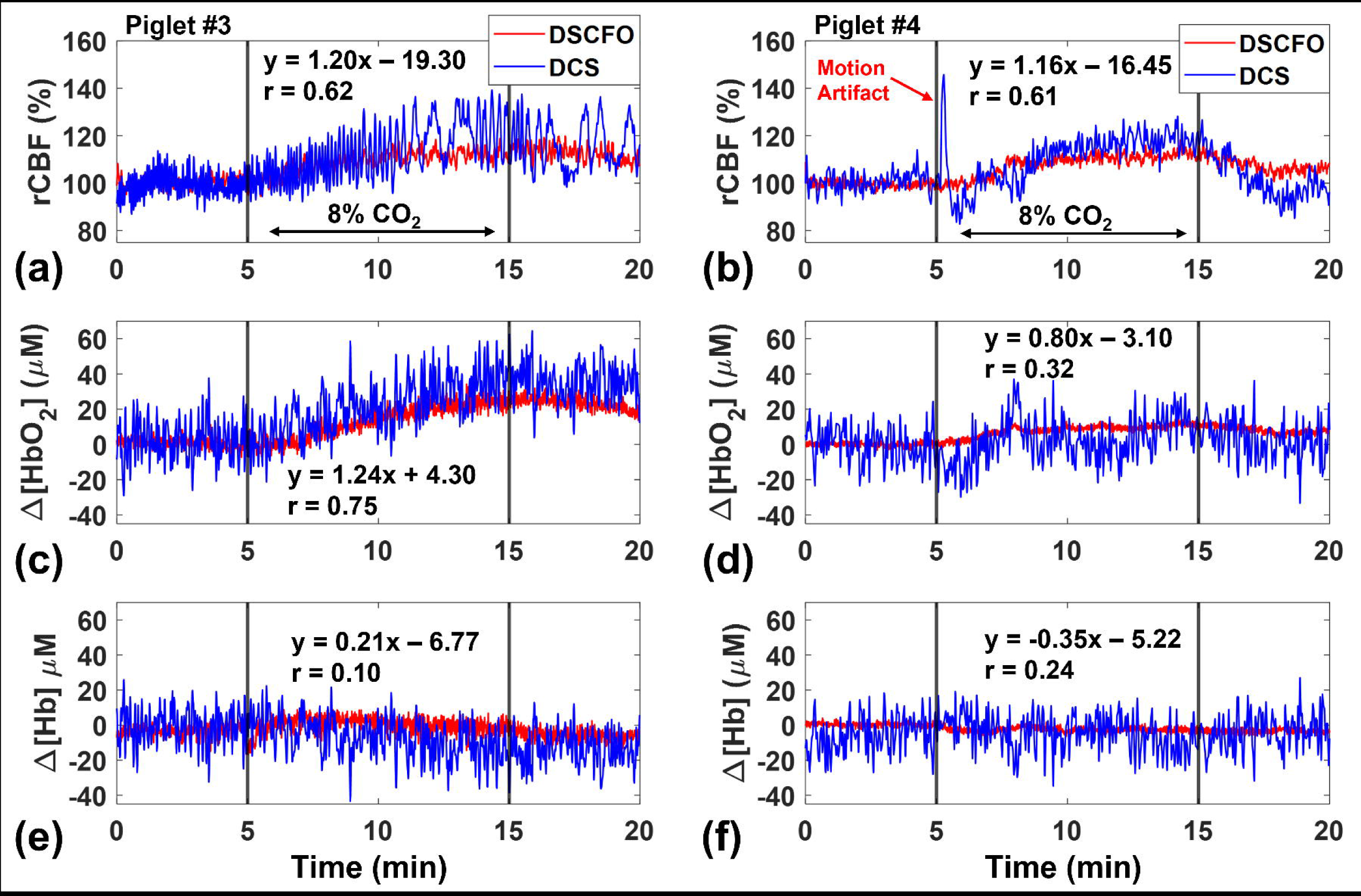
Representative results from Infant #1 and Infant #3 (1^st^ measurement). **(a)** and **(b)** Time-course changes in SpO_2_, HR, RR, rCBF, Δ[HbO_2_], Δ[Hb] from Infant #1 and Infant #3 (1^st^ measurement), respectively. EtCO_2_ was also recorded in Infant #1. The red dashed lines indicate the periods when SpO2 started to change oxygenation desaturation occurred and the red solid lines mark the IH events when SpO_2_ dropped to < 90%, detected by a pulse oximeter.

Pearson correlation coefficients among SpO_2_, HR, RR, rCBF, Δ[HbO_2_], Δ[Hb] were calculated for all 7 measurements for the whole study period (**Table 2**). Note that regional cerebral hemodynamics (rCBF, Δ[HbO_2_] and Δ[Hb]) and peripheral physiological parameters (SpO_2_, HR, and RR) were measured independently by the DSCFO and other commercial devices, respectively. Although no consistent correlation patterns were observed in **Table 2**, these preliminary results from limited number of subjects suggest the importance of multi-parameter measurements for understanding deep insights of peripheral and cerebral regulations.

**Table 2.** Pearson correlation coefficients among SpO_2_, HR, RR, rCBF, Δ[HbO_2_], and Δ[Hb] for 7 measurements from 5 preterm infants.

|  | *Infant #1* | *Infant #2* | *Infant #3, 1<sup>st</sup>* | *Infant #3, 2<sup>nd</sup>* | *Infant #3, 3<sup>rd</sup>* | *Infant #4* | *Infant #5* |
| --- | --- | --- | --- | --- | --- | --- | --- |
| *rCBF/SpO<sub>2</sub>* | 0.02 | -0.02 | 0.07 | 0.63* | -0.40* | 0.16 | -0.16 |
| *rCBF/HR* | -0.17 | -0.02 | -0.48* | -0.52* | -0.25* | -0.19 | -0.30* |
| *rCBF/RR* | -0.05 | -0.05 | 0.20 | 0.22 | -0.03 | 0.23 | 0.46* |
| *Δ[HbO<sub>2</sub>]/rCBF* | 0.35* | -0.30* | -0.54* | 0.53* | 0.04 | 0.01 | 0.46* |
| *Δ[HbO<sub>2</sub>]/SpO<sub>2</sub>* | 0.35* | 0.25 | 0.54* | 0.28* | 0.16 | -0.12 | -0.24 |
| *Δ[HbO<sub>2</sub>]/HR* | -0.23 | 0.03 | 0.58* | -0.10 | -0.01 | 0.26* | -0.42* |
| *Δ[HbO<sub>2</sub>]/RR* | -0.48* | -0.20 | -0.34* | -0.22 | -0.1 | -0.12 | 0.26* |
| *Δ[Hb]/rCBF* | -0.79* | 0.23 | -0.27* | -0.70* | -0.33* | 0.26* | -0.78* |
| *Δ[Hb]/SpO<sub>2</sub>* | -0.22 | -0.36* | 0.54* | -0.43* | 0.15 | 0.14 | 0.07 |
| *Δ[Hb]/HR* | 0.33* | 0.20 | 0.36* | 0.24 | 0.12 | 0.26 | 0.56* |
| *Δ[Hb]/RR* | 0.29* | 0.11 | 0.28* | 0.12 | 0.26* | 0.31 | -0.57* |

Note: “\*” indicate that significant correlations ( $p < 0.05$ ) were observed between two measured variables over the whole study period.

## 4. Discussion

### 4.1 Study motivation and innovation

There is an unmet need for continuous cerebral monitoring at the bedside in NICUs for timely diagnosis and day-to-day management of critically ill preterm infants at risk for brain injury. Portable NIRS/DCS technologies have been utilized for continuous monitoring of cerebral blood oxygenation/CBF variations in preterm infants ^39,40^. However, most systems employ rigid fiber-optic probes for light delivering and detection, which constrain the subject’s movement. By contrast, our innovative DSCFO technique offers advanced features over conventional NIRS/DCS technologies. These advanced features include wearable fiber-free probe, low-cost ergonomic device, rapid data acquisition, real-time data analysis, and multi-parameter measurements (rCBF, Δ[HbO_2_], and Δ[Hb]). 3D-printed wearable DSCFO probes with skin-safe flexible filaments simplify the probe installation on the fragile head of neonatal subjects. Moreover, the thermometer embedded in the DSCFO probe continuously monitors skin temperature, thus ensuring measurement safety. Finally, we have designed unique translational experiments to validate the DSCFO against the established DCS in neonatal piglets and demonstrate the feasibility of DSCFO for continuous cerebral monitoring in preterm infants.

### 4.2 Validation of DSCFO against DCS in neonatal piglets

In Piglet #1 with intact skull, we continuously monitor rCBF during transient global ischemia and asphyxia (**Fig. 2b**) to compare measurement sensitivities of NanEye cameras in the DSCFO and APDs in the DCS. For fair comparisons, both DSCFO and DCS use the same laser source (785 nm long-coherence laser, CrystaLaser) but different detectors (APDs and NanEye cameras). Significant strong correlations between the two measurements were observed, indicating the agreement of DSCFO and DCS in measuring rCBF (**Fig. 3**). The slopes of linear regressions varied from 1.23 to 0.88 (**Fig. 3a-3d**), indicating the difference in detection sensitivity of the two optical sensors. These results agreed with our previous findings in concurrent DCS/DSCF measurement of rCBF in anesthetized mice ^33,34^. Generally, the expensive single-photon-counting APD has a better sensitivity than the low-cost NanEye camera. However, other factors may also impact the measurement, including tissue heterogenous response, probe location, detector window size, and methodology difference ^34,35^.

We noted large spikes in rCBF detected by APDs after right CCA ligation in both hemispheres (⍰ in **Fig. 3a** and **Fig. 3b**). This might be attributed to activation of sympathetic nervous system, leading to an immediate increase in rCBF through contralateral carotid artery to compensate for the loss of rCBF in the ipsilateral hemisphere via the Circle of Willis ^38^. However, these spikes were not captured by the NanEye camera, indicating the difference in detection sensitivities of the two technologies. DCS utilizes a single-mode fiber (∼5 μm) connected to the high-sensitive APD to detect temporal fluctuation of diffuse laser speckles whereas DSCFO uses fiber-free, less-sensitive 2D NanEye camera to detect spatial fluctuation of diffuse laser speckles ^32–35^.

In Piglet #2 with intact head, we compared performances of single-wavelength DSCFO and single-wavelength DCS for continuous monitoring of rCBF during hypercapnia (8%CO_2_) and transient global ischemia (**Fig. 2c**). Similarly, significant strong correlations between the two measurements were observed (**Fig. 4a**). The slope of linear regression was greater than 1 (1.31 in **Fig. 4a**), indicating a better sensitivity of DCS (long-coherence laser plus high-sensitive APD) over DSCFO in detecting rCBF.

In Piglet #3 and Piglet #4 with intact head, we compared effectiveness of dual-wavelength DSCFO and dual-wavelength DCS for continuous monitoring of rCBF, Δ[HbO_2_], and Δ[Hb] during hypercapnia (8%CO_2_) (**Fig. 2d**). Moderate or strong correlations between the two measurements of rCBF and Δ[HbO_2_] were observed (**Fig. 5a-5d**). The slopes of linear regressions in rCBF were also greater than 1 (1.20 in **Fig. 5a** and 1.16 in **Fig. 5b**) while the slopes of linear regressions in Δ[HbO_2_] varied (1.24 in **Fig. 5c** and 0.80 in **Fig. 5d**).

We observed less changes in cerebral oxygenation (especially Δ[Hb]) than in rCBF during hypercapnia (8%CO_2_ inhalation), which are consistent with previous findings ^41,42^. In addition, larger variations were observed from DCS measurements with the fiber-optic probes, which are sensitive to motion artifacts due to hypercapnia-induced potentiation of respiratory activity and animal body trembling. Taken together, smaller changes in Δ[HbO_2_] and Δ[Hb] during hypercapnia with larger motion artifacts in DCS measurements led to weaker correlations between the two measurements of Δ[HbO_2_] and Δ[Hb], compared to rCBF measurements.

### 4.3 Feasibility of DSCFO for continuous brain monitoring in preterm infants

After the translational assessment in neonatal piglets, we further tested the DSCFO for continuous monitoring of rCBF, Δ[HbO_2_], and Δ[Hb] in preterm infants in the NICU. Seven measurements were successfully conducted in 5 preterm infants via placing a flexible and skin-safe wearable DSCFO probe on the head (**Table 1**). Our DSCFO device detected variations in rCBF, Δ[HbO_2_], and Δ[Hb] in preterm infants during IH/cardiorespiratory events (**Fig. 6** and **Supplementary Fig. 2-6**). rCBF increases following the majority of IH events were likely to compensate for global oxygenation loss. Changes in cerebral oxygenation post-IH events varied, likely depending on the severity, duration, and frequency of IH events and changes in rCBF.

No consistent correlation patterns were observed among peripheral and cerebral monitoring parameters (**Table 2**, **Fig. 6**, and **Supplementary Fig. 2-6**). This inconsistency demonstrates the importance of developing tools for direct brain hemodynamics monitoring. We currently do not fully understand the threshold beyond which peripheral hemodynamic changes, such as IH, start to affect cerebral blood flow and oxygenation. The clinical team’s day-to-day management of hemodynamically unstable preterm infants (e.g., adjusting oxygen supplementation, vasopressor support, etc.) will likely benefit from a tool that directly and continuously monitors brain hemodynamics. It is not possible to draw conclusions about the threshold of IH associated with changes in rCBF, Δ[HbO_2_], and Δ[Hb] from this pilot study. However, this study demonstrates the feasibility and safety of continuous cerebral measurements using our wearable DSCFO sensors in the NICU. Results from this study suggest the importance of continuous, longitudinal, and multiparametric measurements for better understanding of long-term impact of IH events on neonatal brains.

## 5. Conclusions

We have developed, optimized, and evaluated an innovative, low-cost, fast, wearable, fiber-free, DSCFO technique for continuous monitoring of rCBF, Δ[HbO_2_], and Δ[Hb] in neonatal piglets and human preterm infants. The DSCFO was first validated against the standard DCS in neonatal piglets with varied physiological/pathological conditions and then tested in preterm infants during IH events in the NICU. Consistent results between the DSCFO and DCS measurements in neonatal piglets were observed. Results from the pilot clinical study with limited number of subjects demonstrated the feasibility of our DSCFO technology to serve as a low-cost wearable sensor for continuous monitoring of multiple cerebral hemodynamic parameters. Future studies will monitor more subjects longitudinally and integrate/correlate multiple cerebral functional parameters with clinical outcomes to identify biomarkers for prediction and management of IH associated brain injury.

## Data Availability Statement

The data that support the findings of this study are available from the corresponding author upon reasonable request.

## Data Availability

All data produced in the present study are available upon reasonable request to the authors

## Acknowledgements

We acknowledge Kimberly Quire and Sara Butler, Department of Pediatrics, University of Kentucky for supporting measurements in NICUs at the Kentucky Children’s Hospital. We also acknowledge Hollie Y. van Rooyen and Jason B. Oakes from the Division of Laboratory Animal Resources at the University of Kentucky for supporting measurements in neonatal piglets.

## Funding

Supported partially by the National Institute of Health (NIH, R01-EB028792 to G. Y.; R01-HD101508 to G. Y.; #R56-NS117587 to G. Y.; R21-HD091118 to G. Y.; and UL1-TR001998 to E. G. A) and University of Kentucky Halcomb Fellowship in Medicine and Engineering to X. L.

## Author Contribution

G.Y., Lei C., E.G.A, H.S.B, and X.L. conceived the study. X.L. designed and built the wearable sensor. X.L., M.M., F.F., S.R.H, P.S. acquired data. Lei C. conducted and supervised animal experiments. E.G.A conducted and supervised human experiments. X.L., Li C., and J.C. analyzed and interpreted data. Li C. performed statistical analysis. Lei C. provided resources for animal experiments. E.G.A and H.S.B provided resources for human experiments. X.L. drafted the manuscript. M.M., F.F., S.R.H, P.S., G.Y., Lei C., E.G.A., H.S.B., Li C., J.C., and X.L. reviewed and edited the manuscript. All authors gave final approval of the version to be published.

## Competing Interests

The authors declare no competing interest.

## Consent Statement

Under a protocol approved by the Institutional Review Board of the University of Kentucky, informed consent was obtained by a physician researcher from each subject’s parents.

---

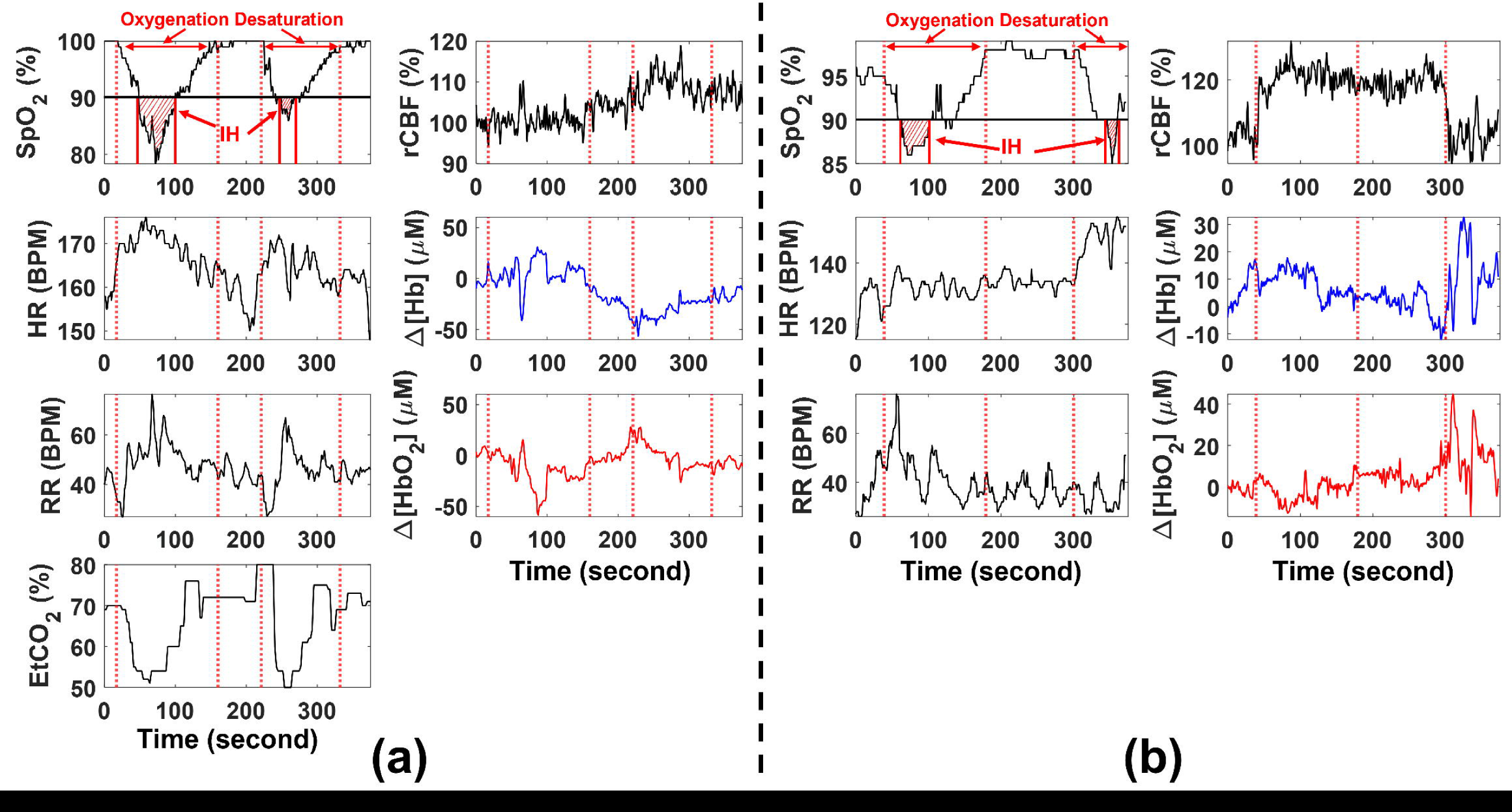

## Notes

### Competing Interest Statement

The authors have declared no competing interest.

### Funding Statement

This study was funded by the National Institute of Health (NIH, R01-EB028792 to G. Y.; R01-HD101508 to G. Y.; #R56-NS117587 to G. Y.; R21-HD091118 to G. Y.; and UL1-TR001998 to E. G. A) and the University of Kentucky Halcomb Fellowship in Medicine and Engineering to X. L.

### Author Declarations

The Institutional Review Board of the University of Kentucky gave ethical approval for this work.

## References

1 Perin, J. et al. Global, regional, and national causes of under-5 mortality in 2000–19: an updated systematic analysis with implications for the Sustainable Development Goals. The Lancet Child & Adolescent Health 6, 106–115 (2022).

2 Poets, C. F. Intermittent hypoxia and long-term neurological outcome: How are they related? Seminars in Fetal and Neonatal Medicine 25, 101072 (2020). 10.1016/j.siny.2019.101072

3 Abu Jawdeh, E. G. Intermittent Hypoxemia in Preterm Infants: Etiology and Clinical Relevance. NeoReviews 18, e637–e646 (2017). 10.1542/neo.18-11-e637

4 Abu Jawdeh, E. G., et al. Intermittent Hypoxemia in Preterm Infants: A Potential Proinflammatory Process. Am J Perinatol 38, 1313–1319 (2020). 10.1055/s-0040-1712951

5 Raffay, T. M. et al. Neonatal intermittent hypoxemia events are associated with diagnosis of bronchopulmonary dysplasia at 36 weeks postmenstrual age. Pediatric Research 85, 318–323 (2019). 10.1038/s41390-018-0253-z

6 Neubauer, J. A. Invited Review: Physiological and pathophysiological responses to intermittent hypoxia. Journal of Applied Physiology 90, 1593–1599 (2001). 10.1152/jappl.2001.90.4.1593

7 Chen, L. et al. Oxidative Stress and Left Ventricular Function with Chronic Intermittent Hypoxia in Rats. American Journal of Respiratory and Critical Care Medicine 172, 915–920 (2005). 10.1164/rccm.200504-560OC

8 Wong, F. Y. et al. Impaired Autoregulation in Preterm Infants Identified by Using Spatially Resolved Spectroscopy. Pediatrics 121, e604–e611 (2008). 10.1542/peds.2007-1487

9 van Bel, F. & Mintzer, J. P. Monitoring cerebral oxygenation of the immature brain: a neuroprotective strategy? Pediatric Research 84, 159–164 (2018). 10.1038/s41390-018-0026-8

10 Verhagen, E. A. et al. Cerebral oxygenation is associated with neurodevelopmental outcome of preterm children at age 2 to 3 years. Developmental Medicine & Child Neurology 57, 449–455 (2015). 10.1111/dmcn.12622

11 Di Fiore, J. M., MacFarlane, P. M. & Martin, R. J. Intermittent Hypoxemia in Preterm Infants. Clin Perinatol 46, 553–565 (2019). 10.1016/j.clp.2019.05.006

12 Martin, R. J., Di Fiore, J. M., MacFarlane, P. M. & Wilson, C. G. in Arterial Chemoreception. (eds Colin A. Nurse, Constancio Gonzalez, Chris Peers, & Nanduri Prabhakar) 351-358 (Springer Netherlands).

13 Fantini, S., Sassaroli, A., Tgavalekos, K. T. & Kornbluth, J. Cerebral blood flow and autoregulation: current measurement techniques and prospects for noninvasive optical methods. Neurophotonics 3, 031411 (2016).

14 Kooi, E. M. W. et al. Measuring cerebrovascular autoregulation in preterm infants using near-infrared spectroscopy: an overview of the literature. Expert Review of Neurotherapeutics 17, 801–818 (2017). 10.1080/14737175.2017.1346472

15 Hyttel-Sorensen, S. et al. Cerebral near infrared spectroscopy oximetry in extremely preterm infants: phase II randomised clinical trial. BMJ (Clinical research ed.) 350, g7635 (2015). 10.1136/bmj.g7635

16 Diop, M., Kishimoto, J., Toronov, V., Lee, D. S. & St Lawrence, K. Development of a combined broadband near-infrared and diffusion correlation system for monitoring cerebral blood flow and oxidative metabolism in preterm infants. Biomedical optics express 6, 3907–3918 (2015). 10.1364/boe.6.003907

17 Andresen, B. et al. Cerebral oxygenation and blood flow in normal term infants at rest measured by a hybrid near-infrared device (BabyLux). Pediatric Research 86, 515–521 (2019). 10.1038/s41390-019-0474-9

18 Wyser, D., Lambercy, O., Scholkmann, F., Wolf, M. & Gassert, R. J. N. Wearable and modular functional near-infrared spectroscopy instrument with multidistance measurements at four wavelengths. 4, 041413–041413 (2017).

19 Lacerenza, M. et al. Wearable and wireless time-domain near-infrared spectroscopy system for brain and muscle hemodynamic monitoring. Biomedical optics express 11, 5934–5949 (2020). 10.1364/BOE.403327

20 Funane, T. et al. Rearrangeable and exchangeable optical module with system-on-chip for wearable functional near-infrared spectroscopy system. 5, 011007–011007 (2018).

21 Roche-Labarbe, N. et al. Noninvasive optical measures of CBV, StO2, CBF index, and rCMRO2 in human premature neonates’ brains in the first six weeks of life. 31, 341-352 (2010). 10.1002/hbm.20868

22 Buckley, E. M. et al. Cerebral hemodynamics in preterm infants during positional intervention measured with diffuse correlation spectroscopy and transcranial Doppler ultrasound. Opt. Express 17, 12571–12581 (2009). 10.1364/OE.17.012571

23 Shang, Y. et al. Cerebral monitoring during carotid endarterectomy using near-infrared diffuse optical spectroscopies and electroencephalogram. Physics in Medicine & Biology 56, 3015 (2011). 10.1088/0031-9155/56/10/008

24 Cheng, R., Shang, Y., Hayes, D., Saha, S. P. & Yu, G. Noninvasive optical evaluation of spontaneous low frequency oscillations in cerebral hemodynamics. NeuroImage 62, 1445–1454 (2012). 10.1016/j.neuroimage.2012.05.069

25 Sunwoo, J. et al. Diffuse correlation spectroscopy blood flow monitoring for intraventricular hemorrhage vulnerability in extremely low gestational age newborns. Scientific Reports 12, 12798 (2022). 10.1038/s41598-022-16499-3

26 Farzam, P. et al. Shedding light on the neonatal brain: probing cerebral hemodynamics by diffuse optical spectroscopic methods. Scientific Reports 7, 15786 (2017). 10.1038/s41598-017-15995-1

27 Shang, Y. et al. Portable optical tissue flow oximeter based on diffuse correlation spectroscopy. Opt. Lett. 34, 3556–3558 (2009). 10.1364/OL.34.003556

28 White, B. R., Liao, S. M., Ferradal, S. L., Inder, T. E. & Culver, J. P. Bedside optical imaging of occipital resting-state functional connectivity in neonates. Neuroimage 59, 2529–2538 (2012). 10.1016/j.neuroimage.2011.08.094

29 Frijia, E. M. et al. Functional imaging of the developing brain with wearable high-density diffuse optical tomography: A new benchmark for infant neuroimaging outside the scanner environment. Neuroimage 225, 117490 (2021). 10.1016/j.neuroimage.2020.117490

30 Selb, J. et al. Prolonged monitoring of cerebral blood flow and autoregulation with diffuse correlation spectroscopy in neurocritical care patients. Neurophotonics 5, 045005 (2018). 10.1117/1.NPh.5.4.045005

31 Liu, X. et al. Simultaneous measurements of tissue blood flow and oxygenation using a wearable fiber-free optical sensor. Journal of Biomedical Optics 26, 012705 (2021).

32 Liu, X. et al. in Biophotonics in Exercise Science, Sports Medicine, Health Monitoring Technologies, and Wearables III. PC1195601 (SPIE).

33 Liu, X. et al. in Biophotonics in Exercise Science, Sports Medicine, Health Monitoring Technologies, and Wearables II. 116380A (SPIE).

34 Liu, X. et al. A Wearable Fiber-free Optical Sensor for Continuous Monitoring of Cerebral Blood Flow in Freely Behaving Mice. IEEE Transactions on Biomedical Engineering, 1–11 (2022). 10.1109/TBME.2022.3229513

35 Huang, C. et al. A wearable fiberless optical sensor for continuous monitoring of cerebral blood flow in mice. IEEE J Sel Top Quantum Electron 25 (2019). 10.1109/JSTQE.2018.2854597

36 Huang, C. et al. Speckle contrast diffuse correlation tomography of cerebral blood flow in perinatal disease model of neonatal piglets. J Biophotonics 14, e202000366 (2021). 10.1002/jbio.202000366

37 Huang, C. et al. Noninvasive noncontact speckle contrast diffuse correlation tomography of cerebral blood flow in rats. Neuroimage 198, 160–169 (2019).

38 Vrselja, Z., Brkic, H., Mrdenovic, S., Radic, R. & Curic, G. Function of Circle of Willis. 34, 578–584 (2014). 10.1038/jcbfm.2014.7

39 Kebaya, L. M. N. et al. Three-dimensional cranial ultrasound and functional near-infrared spectroscopy for bedside monitoring of intraventricular hemorrhage in preterm neonates. Scientific Reports 13, 3730 (2023). 10.1038/s41598-023-30743-4

40. Otic, N. et al. in Biophotonics Congress: Biomedical Optics 2022 (Translational, Microscopy, OCT, OTS, BRAIN). JM3A.70 (Optica Publishing Group).

41 Huppert, T. J. et al. Sensitivity of neural-hemodynamic coupling to alterations in cerebral blood flow during hypercapnia. 14, 044038–044038-044016 (2009).

42 Rostrup, E., Law, I., Pott, F., Ide, K. & Knudsen, G. M. Cerebral hemodynamics measured with simultaneous PET and near-infrared spectroscopy in humans. Brain Research 954, 183–193 (2002). 10.1016/S0006-8993(02)03246-8

