## Supplementary Materials for "A Wearable Fiber-Free Optical Sensor for Continuous Monitoring of Neonatal Cerebral Blood Flow and Oxygenation"

**Wearable Fiber-free DSCFO System**

Details about the DSCFO technology for use in small rodents and human forearm muscles are found in our previous publications [31-35]. **Supplementary Fig. 1a-1d** show the modified DSCFO system with an upscaled wearable probe designed particularly for continuously monitoring CBF and cerebral oxygenation in preterm infants. Briefly, a compact DSCFO controlling unit ② (**Supplementary Fig. 1b**) was formed by stacking a miniaturized current driver PCB ③ (**Supplementary Fig. 1c**) with a microcontroller (Arduino Uno, SparkFun) ④ and a camera controlling board (NanoUSB 2.2, Awaiba) ⑤. An ultra-small low-power camera ⑦ (NanEye 2D Black and White; dimension: 1 mm × 1 mm, power: 4 mW, Awaiba) was used as a 2D detector array to provide a 250 × 250 pixel-array (pixel dimensions: 3 µm × 3 µm). Two small laser diodes ⑧ (L785P25, Ø5.6 mm, 30 mW, 785 nm; HL8338MG, Ø5.6 mm, 50 mW, 830 nm, Thorlabs) were electrically connected to and powered by the driving circuit ③ (dimension: 44 mm × 79 mm) which embedded a light power stabilization module, as was done in previous DSCFO device [31]. A built-in photodiode in the laser diode package continuously detects the light intensity generated by the laser diode and generates the feedback current proportionally. Arduino Uno reads this feedback current to stabilize light intensity output.

The entire controller assembly (i.e., current driver, Arduino, and camera boards) was housed within an electrically insulated box and powered/directed by a laptop ①. The DSCFO controller and laptop communications went through two USB cables. A graphic user interface (**Supplementary Fig. 1b**) was developed using Microsoft C# to control the camera and display the results in real time. A LabVIEW (National Instruments) program was designed to monitor and control light intensities of laser diodes. Synchronization between the camera and the laser diodes was accomplished using the internal control protocol communication (i.e., C# to LabVIEW) through the laptop lookback address. The raw intensity images (**Fig. 1a**), light intensity, and calculated cerebral blood flow and oxygenation information were displayed on the screen in real time. All raw images were stored on the local drive for further off-line data analysis.

The NanEye camera, two laser diodes, and a thermometer T (Cryo Temp, CR-1) were affixed to the upscaled wearable DSCFO probe ⑥ and connect to the compact device ② through flexible electrical wires. This new DSCFO probe was fabricated by a 3D printer (Qidi X-Max) with soft, flexible, and skin-safe filaments (Chinchilla™ flexible 3D printer filament, NinjaTek) and then gently attached on the forehead of a sleeping preterm infant using the extended flexible bands (**Supplementary Fig. 1d**). The soft probe and its extended bands were then covered by a medical elastic band to prevent ambient light influence. Note that the two laser diodes were installed ~2 mm above the skin to ensure sufficient dissipation of the heat generated by the laser diodes. A thermometer ⑨ was placed close to the laser diodes for continuous monitoring of skin temperature. A pulse oximeter ⑩ was used to measure SpO_2_ continuously.

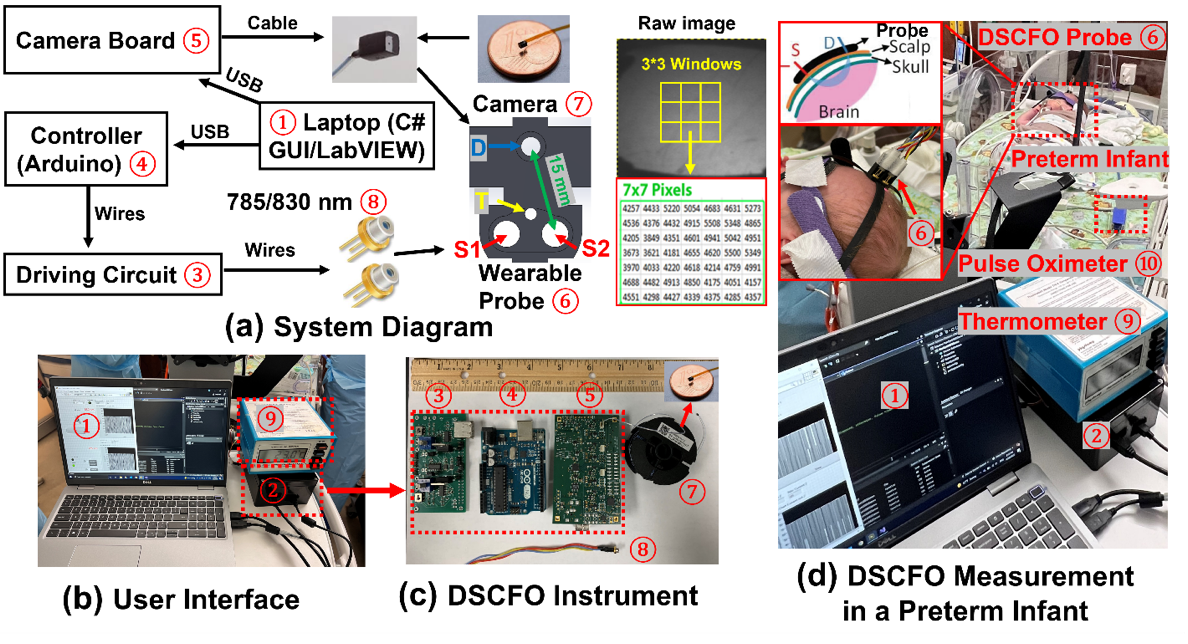

**Supplementary Figure 1:** The DSCFO system for continuous cerebral monitoring of human preterm infants in the NICU. **(a)** System diagram. **(b)** User Interface ①, DSCFO device ②, and a thermometer ⑨. **(c)** Components for DSCFO device including customized circuit ③ to drive laser diodes ⑧, Arduino Uno microcontroller ④, camera electronic (NanEye USB 2.0) ⑤, DSCFO probe ⑥, and NanEye camera ⑦. **(d)** DSCFO measurement in a preterm infant. A thermometer ⑨ was used to continuously detect skin temperature for safety. A pulse oximeter ⑩ was used to continuously measure SpO_2_. The DSCFO probe dimensions were 25 × 25 × 20 mm^3^. The source-detector (S-D) distances were 15 mm.

**Fiber-optic DCS System**

The dual-wavelength DCS device was built in-house with two long-coherence lasers at 785 nm and 852 nm (DL785-100-S, DL852-100-S, CrystaLaser) as point sources and four single-photon-counting avalanche photodiodes (APDs, Pacer) as multichannel detectors connected to a four-channel autocorrelator board (Correlator.com). The lasers were coupled to multimode fibers (core diameter: 200 μm) and APDs connected to single-mode fibers (core diameter: 5.6 μm). The source and detector fibers were confined by a black foam pad to form a fiber-optic probe. Like the DSCFO, photons emitted from the laser sources travel through the measured tissue volume via a banana shape pathway and only some of them are detected by the APDs. The APD signals were fed into the autocorrelator yielding autocorrelation curves in parallel every 44 ms. Multiple autocorrelation curves were averaged in ~0.5 seconds (equivalent sampling rate: 2 Hz) to improve the SNR. BFI information was extracted by fitting the averaged autocorrelation curve whose decay rate depended mainly on the motion of red blood cells [27, 43]. rCBF was reported for comparisons with those obtained by the DSCFO. Blood oxygenation changes (Δ[HbO_2_] and Δ[Hb]) were obtained from the light intensity variations measured at the two wavelengths (785 nm and 852 nm) using the modified Beer-Lambert law [44].

**Results from Preterm Infants**

**Supplementary Figure 2** shows results from Infant #2. Two IH events occurred with SpO_2_ dropped to less than 80%. There was a HR deceleration following both IH events. RR fluctuated but predominantly increased during the IH events. rCBF increased while Δ[HbO_2_] decreased and Δ[Hb] increased following the first IH event and fluctuated during the second IH event.

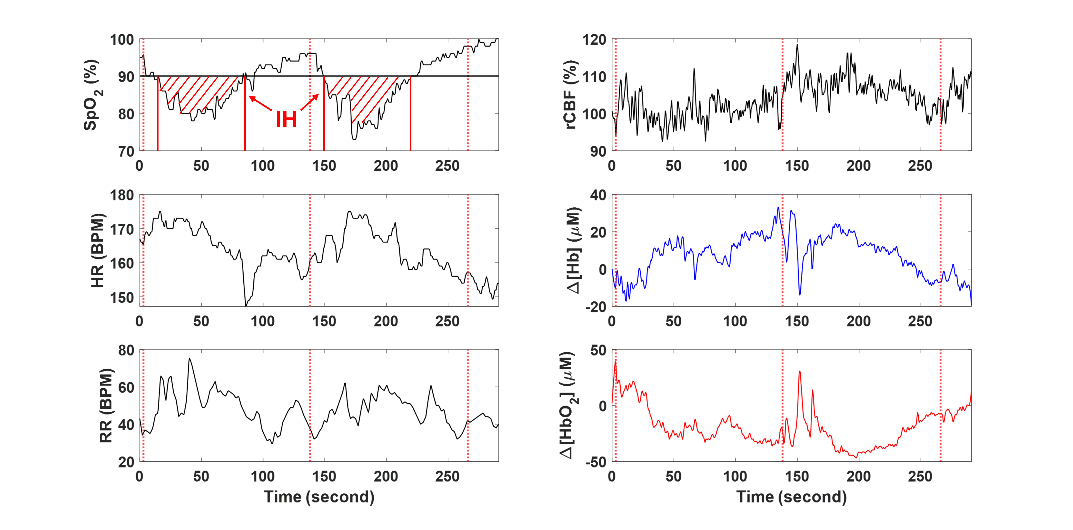

**Supplementary Figure 2:** Measurement results from Infant #2.

**Supplementary Figure 3** shows results of the second measurement from Infant #3. Two IH events recorded with SpO_2_ dropped to less than 80%. There was an initial increase in HR followed by a deceleration after the first IH event. RR fluctuated during the first IH event with no apparently significant pattern. DSCFO recorded an associated decrease in rCBF but relatively stable cerebral oxygenation.

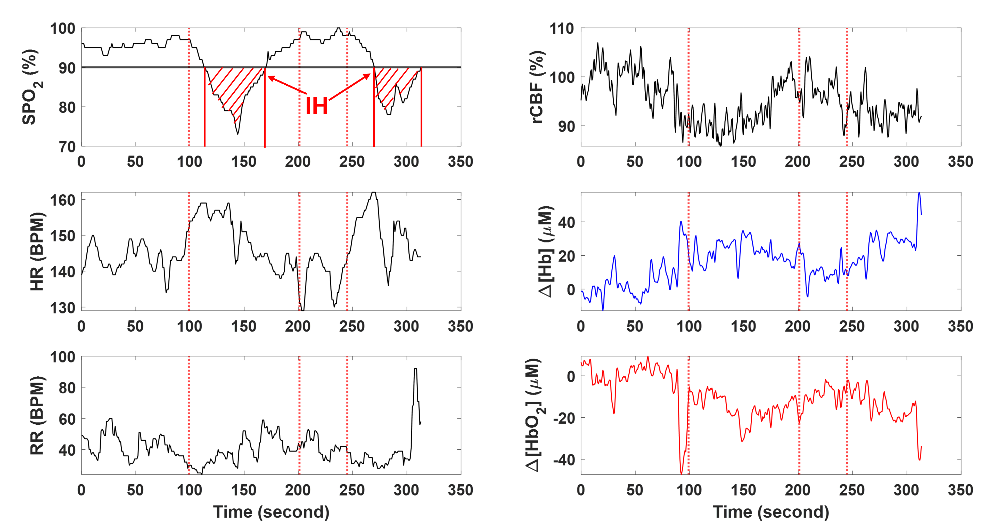

**Supplementary Figure 3:** Measurement results from Infant #3 during the second measurement.

**Supplementary Figure 4** shows results of third measurement from Infant #3. This patient presented a prolonged IH event associated with the deceleration in HR. In response, rCBF increased and Δ[HbO_2_] decreased. At the end of the study period, there was a second brief IH event that was not associated with significant changes in cerebral blood flow and oxygenation. RR fluctuated during the measurement.

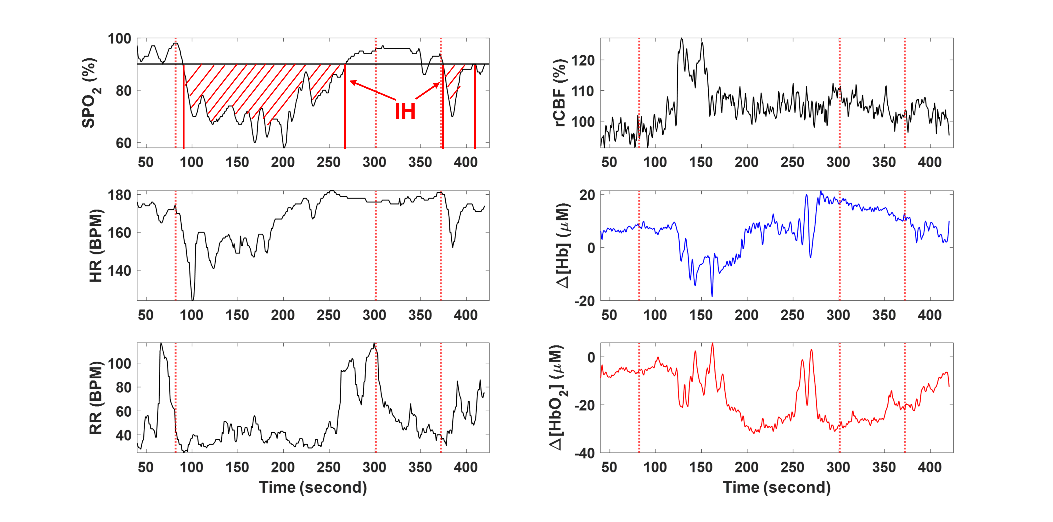

**Supplementary Figure 4:** Measurement results from Infant #3 during third measurement.

**Supplementary Figure 5** shows results from Infant #4. There are two brief IH events associated with a decrease in HR and an increase in RR. rCBF increased post IH events along with a decrease in Δ[HbO_2_] and an increase in Δ[Hb].

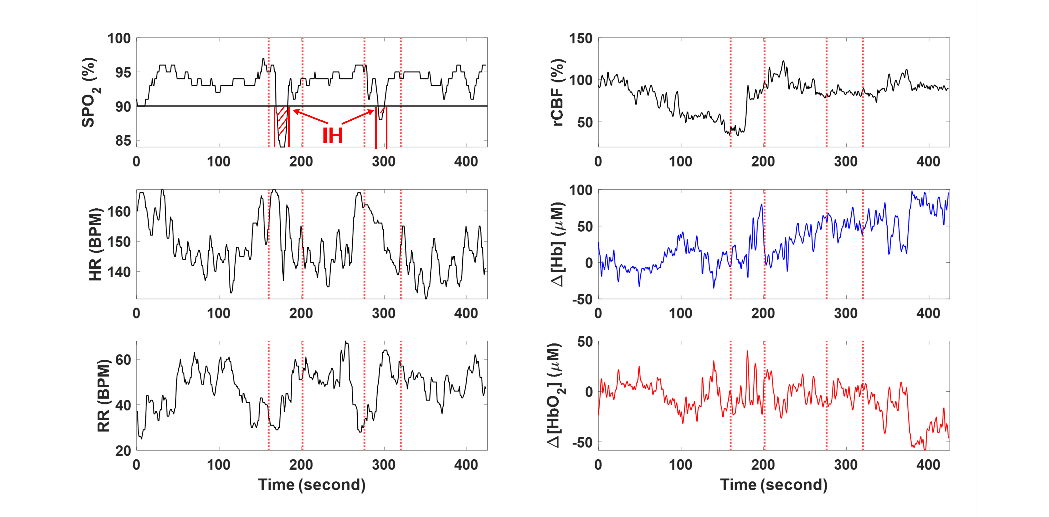

**Supplementary Figure 5:** Measurement results from Infant #4.

**Supplementary Figure 6** shows results from Infant #5. Two IH events were recorded. RR initially decreased followed by a periodic breathing pattern until the end of study period. There were minor fluctuations in HR. Following the first IH event, rCBF and Δ[Hb] increased while Δ[HbO_2_] slightly decreased. No notable changes occurred after the second IH event.

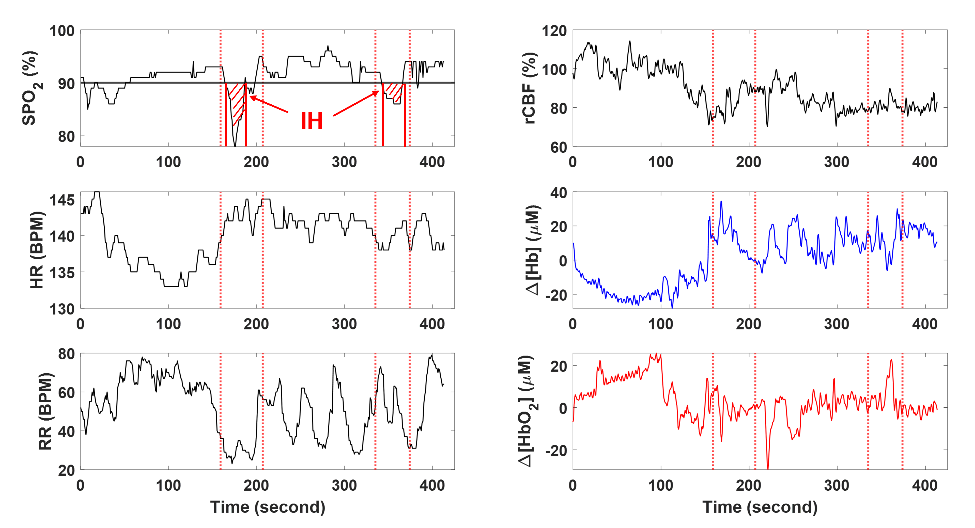

**Supplementary Figure 6:** Measurement results from Infant #5.
